# Detecting impaired language processing in MCI patients using around-the-ear cEEgrid electrodes

**DOI:** 10.1101/2021.03.18.21253911

**Authors:** K. Segaert, C. Poulisse, R. Markiewicz, L. Wheeldon, D. Marchment, Z. Adler, D. Howett, D. Chan, A. Mazaheri

## Abstract

Mild cognitive impairment (MCI) is the term used to identify those individuals with subjective and objective cognitive decline but with preserved activities of daily living and an absence of dementia. While MCI can impact functioning in different cognitive domains, most notably episodic memory, relatively little is known about the comprehension of language in MCI. In this study we used around-the-ear electrodes (cEEGrids) to identify impairments during language comprehension in MCI patients. In a group of 23 MCI patients and 23 age-matched controls, language comprehension was tested in a two-word phrase paradigm. We examined the oscillatory changes following word onset as a function of lexico-semantic single-word retrieval (e.g. *swrfeq* versus *swift*) and multi-word binding processes (e.g. *horse* preceded by *swift* versus preceded by *swrfeq*). Electrophysiological signatures (as measured by the cEEGrids) were significantly different between MCI patients and controls. In controls, lexical retrieval was associated with a rebound in the alpha/beta range and binding was associated with a post-word alpha/beta suppression. In contrast, both the single-word retrieval and multi-word binding signatures were absent in the MCI group. The signatures observed using cEEGrids in controls were comparable to those signatures obtained with a full-cap EEG set-up. Importantly, our findings suggest that MCI patients have impaired electrophysiological signatures for comprehending single-words and multi-word phrases. Moreover, cEEGrids set-ups provide a non-invasive and sensitive clinical tool for detecting early impairments in language comprehension in MCI.

## Introduction

Mild cognitive impairment (MCI) is characterized by cognitive decline, most notably in the domain of episodic memory. Roughly 60% of individuals diagnosed with MCI progress to develop dementia within 5 years of MCI diagnosis (Gauthier et al., 2006; Portet et al., 2006). MCI patients with deficits in multiple cognitive domains are more likely to develop dementia due to Alzheimer’s disease (AD) than MCI patients with impairment in a single cognitive domain (Alexopoulos, Grimmer, Perneczky, Domes, & Kurz, 2006; Bozoki, Giordani, Heidebrink, Berent, & Foster, 2001). This underlines the importance of evaluating other domains beyond traditional memory assessment. Language is a domain of particular relevance given that language impairments occur early in dementia (Caramelli, Mansur, & Nitrini, 1998). As such, sensitive measures that elucidate language deficits in MCI patients can be of great diagnostic and prognostic value. In this study we use a novel, wearable EEG set-up (i.e. cEEGrids (Bleichner & Debener, 2017)) to study language comprehension, at the level of single-words as well as multi-word utterances, in MCI patients.

### Language impairments in MCI patients

Early language deficits in dementia are well described (Caramelli et al., 1998; Henry, Crawford, & Phillips, 2004). In the prodromal stage of AD, subtle impairments may be involved in several cognitive domains, including language. MCI patients constitute a prime target for investigating predictive markers of AD since annual conversion to dementia of MCI is around 10% in specialist clinics and around 5% in the community (Mitchell & Shiri-Feshki, 2009). Moreover, individuals with MCI who have impairments in multiple cognitive domains, including language, are more likely to progress to dementia (Alexopoulos et al., 2006; Bozoki et al., 2001; Taler & Phillips, 2008). As such, there is an interest in being able to detect cognitive deficits in MCI patients early on, including deficits in those domains which might be subtle and more difficult to detect, such as language.

Language impairments have previously been shown in MCI patients (for a comprehensive review see: Taler & Phillips, 2008). In language *production*, there are single-word naming deficits (most notably naming of people and buildings (Ahmed, Arnold, Thompson, Graham, & Hodges, 2008) as well as deficits in the production of discourse (Eyigoz, Mathur, Santamaria, Cecchi, & Naylor, 2020; Roark, Mitchell, Hosom, Hollingshead, & Kaye, 2011). Less is known about the *comprehension* of language, compared to production. Perhaps as a consequence, most of the standardized neuropsychological tests rely on measures of language production only, rather than a combination of production and comprehension (although see Ritchie et al. (2001) for an exception). In this paper we explore whether comprehension deficits for single-words and word-pair combinatorics can be detected in MCI patients.

Those studies that have measured language *comprehension* using behavioral performance measures in MCI patients have generated differing observations. Some have shown that word comprehension (as measured in a lexical decision task and priming task) is affected in dementia but unaffected in MCI (Duong, Whitehead, Hanratty, & Chertkow, 2006), while others have identified impairments in accessing lexical information (also measured using a lexical decision task) in MCI patients compared to controls (Taler & Jarema, 2006). One possible explanation for the discrepant findings is that language comprehension deficits may be difficult to detect by using only behavioural measures. Behavioral performance measures necessarily involve additional decision and response processes which may mask subtle differences in the language comprehension process itself. An alternative is to use methods which allow a real-time investigation as language comprehension unfolds over time, such as EEG (Weiss & Mueller, 2003).

Indeed, a few neuroimaging studies have revealed word comprehension impairments in MCI patients, in the absence of overt behavioural deficits. Functional MRI studies have revealed that MCI patients had reduced activation in the posterior left superior temporal sulcus (STS) compared to healthy controls for word processing (Vandenbulcke, Peeters, Dupont, Van Hecke, & Vandenberghe, 2007). Olichney et al. (2008) examined the electrophysiological response to words in MCI patients and found that modulations in ERP components sensitive to language (the N400 and P600) had diagnostic and prognostic value; those MCI patients characterized by abnormal N400 or P600 word repetition effects had an increased likelihood of conversion to dementia. Mazaheri et al. (2018) reported follow-up analyses of this cohort of MCI patients and focused on the power changes in oscillatory activity induced by the presentation of words. In this study, word presentation induced an early increase in theta (3–5 Hz) activity (i.e. processing of the word form: Bastiaansen, Oostenveld, Jensen, & Hagoort, 2008; Bastiaansen, Van Der Linden, Ter Keurs, Dijkstra, & Hagoort, 2005), followed by alpha (∼10 Hz) suppression (i.e. post-perceptual processing of sensory information (Pfurtscheller, 2001) and allocation of resources according to processing demands (Foxe, Simpson, & Ahlfors, 1998) over posterior sites. The MCI convertors group (i.e. patients that would go on to convert to AD in 3-years’ time) had a diminished early theta power increase induced by presentation of the word compared to MCI non-convertors and controls.

Taken together, two initial observations arise from the limited number of studies conducted in this field to date. First, the detection of language comprehension impairments in MCI patients may be of diagnostic and prognostic value. The sensitivity and specificity of a language comprehension measure might be at least equal to that of other measures for the diagnosis of MCI. A next step furthermore could be to investigate whether the combination of language measures to memory tests would increase sensitivity and specificity of diagnosis and/or prognosis. Second, previous research suggests there are electrophysiological markers that correspond to language impairments in MCI. This work is taken forward in the current study where we investigate the comprehension of single words as well as multi-word phrases. Language comprehension unfolds over time, therefore real-time temporal measures such as EEG might be more sensitive indicators of comprehension impairments than behavioural performance measures, which examine comprehension off-line and typically involve a decision making process.

### C-shaped electrode arrays (cEEGrids) as a measurement tool

In this study we use a novel EEG set-up to study the electrophysiological signatures of language comprehension in MCI patients. We used C-shaped (see Figure 1 for layout) electrode arrays (cEEGrids), placed around the ear (Bleichner & Debener, 2017). The cEEGrids are fast and easy to apply, lightweight and comfortable to wear. They avoid some important drawbacks of patient research using traditional high-density EEG, for which cap-preparation and clean-up is more time-consuming. Auditory processing studies have demonstrated that around-the-ear cEEGrid electrodes can capture cortical electrophysiological potentials and alpha oscillations, with signals that closely correspond to those recorded with full-cap scalp-EEG (Bleichner & Debener, 2017; Bleichner, Mirkovic, & Debener, 2016; Debener, Emkes, De Vos, & Bleichner, 2015). Previous research suggests high suitability of cEEGrids for sleep staging (Sterr et al., 2018) and diagnosis of hearing loss (Garrett, Debener, & Verhulst, 2019), with potential applications using cEEGrids in brain-computer-interface steering of hearing aids (Mirkovic, Bleichner, De Vos, & Debener, 2016). A study focusing on visual and motor processing revealed that cEEGrids are best suited for recording activity from posterior scalp sites (Pacharra, Debener, & Wascher, 2017). This previous work with cEEGrids motivated our proof-of-concept study on language processing in a population of patients with MCI, to establish if there are wider clinical applications for the use of cEEGrids.

**Figure 1.** *Layout of the cEEGrid electrodes.* The right mastoid (R5) served as the ground while the left mastoid (L6) served as the reference during the online recordings. The data were offline re-referenced to a linked mastoid where L6 and R6 served as the reference.

### The present study design

We investigate modulations in oscillatory brain activity that support language comprehension in MCI patients and healthy older adult controls. Our aims are as follows. First, we aim to extend on previous work (Mazaheri et al., 2018; Olichney et al., 2008) revealing altered electrophysiological signatures for single-word processing in MCI patients, by investigating impairments that go beyond the comprehension of single words. The comprehension of phrases and sentences is a key aspect of language processing. Language users construct complex meaning from more elementary semantic building blocks (Hagoort, 2020; Hagoort et al., 2009). The meaning of an individual word (e.g. flat) is altered by the meaning of following words (e.g. flat tyre vs. flat note), such that the combined meaning is greater than that of those words in isolation (Hagoort et al., 2009; Keenan, 1979). This illustrates the unique and expressive power of language: we have the ability to combine words in novel ways to create sentences, which forms the basis for communication and social interactions. The processing which language users need to complete for a two-word phrase forms the foundation of binding in the context of more increasing complexity. The present work investigates elementary binding by means of a minimal two-word phrase paradigm. This offers the advantage of focusing on the binding process while minimizing contributions of other processes involved in language comprehension, such as working memory. Finally, we use a novel cEEGrid set-up and compare the oscillatory modulations as revealed using the cEEGrid set-up with those obtained using a full-cap EEG set-up.

We present two-word phrases that consist of the combination of a non-word with a word (e.g. *swrfeq horse*), as well as two-word phrases for which a combined meaning representation can be formed and binding takes place (e.g. *swift horse*). The comparison of *swrfeq* versus *swift* (i.e. the first word in each condition) yields a signature for single-word retrieval, while the comparison of *horse* preceded by *swrfeq* versus *horse* preceded by *swift* (i.e. the second word in each condition) yields a signature for multi-word binding combinatorics.

More specifically, the electrophysiological signature for single-word retrieval (i.e. the first word in each condition) identifies lexico-semantic retrieval, since retrieval of lexico-semantic properties is possible for real words but not non-words, as well as recognition of the word form and orthographic processing (Taylor et al., 2013).

The electrophysiological signature for multi-word binding combinatorics (i.e. the second word in each condition), identifies a signature for semantic binding, and to some extent, syntactic binding. In both conditions when *horse* is presented, lexico-semantic retrieval takes place. But only in the binding condition (i.e. swift horse) can a complex meaning representation be built for the phrase, based on the elementary building blocks of each individual word. Similar paradigms have been used previously to investigate combinatoris, or, binding processes (Bemis & Pylkkänen, 2013; Pylkkänen et al., 2014; Segaert, et al., 2018; Zaccarella et al., 2017; Zaccarella & Friederici, 2015), including in the ageing literature (Poulisse, Wheeldon, Limachya, Mazaheri, & Segaert, 2020; Poulisse, Wheeldon, & Segaert, 2019).

In summary, we investigate language comprehension impairments in MCI patients. We focus on EEG rather than behavioural performance, since EEG measures the time-course of comprehension as language unfolds and previous research on MCI patients (reviewed above) suggests that this provides a more sensitive measure of language impairment. Language comprehension will be tested in a two-word phrase paradigm to reveal electrophysiological signatures for both *single-word retrieval* of lexico-semantic properties and the computation of multi-word *binding* combinatorics, using cEEGrids given their ease of use. We test whether differential signatures for language comprehension can be detected in MCI patients, focusing on single-word language comprehension (lexical retrieval) as well as the comprehension of multi-word utterances (semantic binding). We examine also whether electrophysiological signatures observed with cEEGrids are comparable to those observed with full-cap EEG set-ups.

## Method

### Participants

27 MCI patients and 27 healthy older adult controls participated in the *cEEGrid* experiment. One patient was excluded because a later MRI revealed an arachnoid cyst, while three patients and four control participants were excluded from the analysis due to extreme noise in the EEG data and signal drop out, resulting in a final sample of 23 MCI patients (mean age: 70 years; SD: 9; range: 51-86 years; 13 males) and 23 healthy older adult controls (mean age: 72 years; SD: 5, range: 61-80 years; 12 males). We should note that these artefacts were not due to the cEEGrids, but due to the custom adapter connecting them to the EEGOSPORTs amplifier. This custom adapter would at times become loose during the recording due to participant movements. For the purpose of comparison between cEEGrid and 64-channel full-cap EEG, we furthermore report the results of a group of 29 healthy older adults (mean age: 73.6 years; SD: 5.8; range: 63-84 years; 13 males) measured on the same paradigm with 64-channel *full-cap EEG* (their data are also reported elsewhere as part of a different study, which focuses on a comparison between young and older adults: Markiewicz et al., submitted).

All participants were right-handed, British English monolingual speakers with normal or corrected-to-normal vision and no diagnosis of dyslexia. All healthy older adults scored in the normal range on the Mini-Mental State Examination (MMSE) or Montreal Cognitive Assessment (MOCA): cEEGrid group: M=29.22, SD: 0.80 on MMSE (O’Bryant et al., 2008); 64-channel full-cap EEG group: M = 27.79, SD = 1.01 on MOCA (Nasreddine et al., 2005). All of the cEEGrid control participants, and 22 of the MCI patients completed the Addenbrooke’s Cognitive Examination Revised (ACE-R) test (Mioshi, Dawson, Mitchell, Arnold, & Hodges, 2006). The control participants had a mean ACE-R of 95/100 (range 83-100), while the MCI patients had a score of 85/100 (range (64-99). The healthy control group had a significantly higher ACE-R score than the MCI patient group (t(1,43)= 4.6300, p<0.0001). There was no significant difference in the number of years spent in education between the groups; cEEGrid healthy controls had an average of 16 years of education (SD: 3), 64-channel full-cap EEG healthy controls had an average of 15 years of education (SD: 3), while the MCI group had an average of 14 years of education (SD: 4).

The MCI patients were recruited from the Cambridge University Hospital NHS Trust MCI and Memory Clinics. MCI was diagnosed by a neurologist according to the Petersen criteria (Petersen, 2004), namely (i) the presence of a complaint of defective memory from the patient (generally corroborated by an informant); (ii) an objective memory impairment for age on formal testing; (iii) relatively preserved general cognition for age; (iv) generally intact activities of daily living; (v) no diagnosis of dementia. Control participants (cEEGrid and full-cap EEG) were recruited via the database of the School of Psychology of Birmingham University and were tested at Birmingham University. Participants signed informed consent, which followed the guidelines of the British Psychology Society code of ethics. Ethical approval was obtained from the NHS Cambridge South Research Ethics Committee; the University of Cambridge Human Biology Research Ethics Committee and by the University of Birmingham Ethical Review (ERN 15-0866).

### Design, materials and task

We used a language comprehension paradigm with two-word-phrases, such as “swift horse” and “swrfeq horse” (Figure 2). The comparison of the first word taps into single-word retrieval (i.e. real words, compared to letter strings). Comparison for the second word taps into binding (words in a binding context, compared to words in a no binding context).

**Figure 2.**
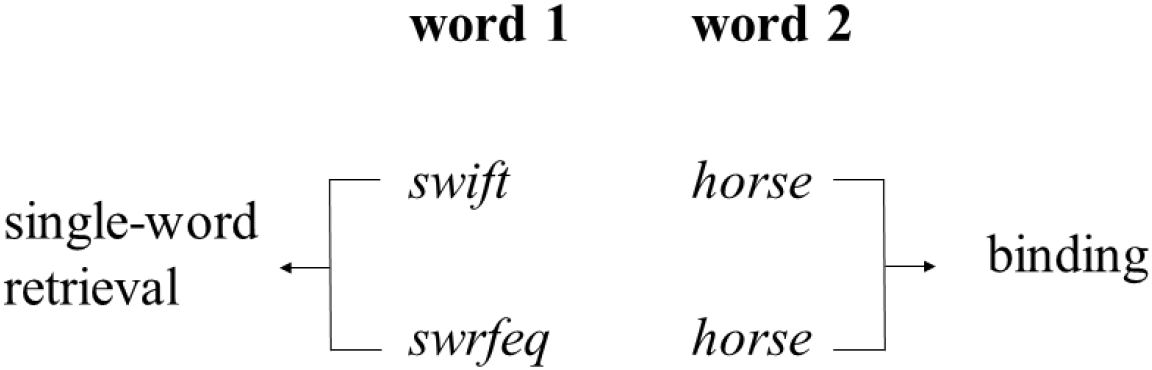
Example stimuli to illustrate the crucial condition contrasts, i.e. for words 1: single-word retrieval; for word 2: binding.

The number of letters between the letter strings and real words were matched (average number of letters in word 1 = 5.8 with sd= 0.76, average number of letters in letter strings = 5.7 with sd = 0.75).

In each condition, about half of the target words were animate and half inanimate. Furthermore, within the binding condition, half of the word pairs were plausible (e.g. swift horse) and the other half were implausible (e.g. barking horse), but for the purpose of the present study these were collapsed. This decision was made because previous analyses of full-cap EEG data on both young and healthy older adults with this same paradigm revealed no differences between the plausible and implausible conditions (Markiewicz, Segaert, & Mazaheri, submitted) suggesting the plausibility manipulation as implemented in this stimulus set may not have been strong enough to yield differences. Also in the present dataset no differences were observed between the plausible and implausible word pairs.

To ensure participants paid attention to the stimuli throughout the experiment, we included yes-no questions about the word pairs on a subset of the trials (22% of all trials). The questions asked “Did you just see [word pair]”. There were no significant differences between the groups in response accuracy (MCI group: mean = 93%, SD: 0.25; healthy older cEEGrid group: mean = 98%, SD: 0.15; healthy older 64-channel full-cap group: mean = 96%, SD = 0.2; all p >.1). Each individual participant scored higher than 80%. Note that these behavioural performance data by no means serve as a sensitive measure of language comprehension performance. Rather, from these behavioural data we can conclude that all participants paid close attention to the stimuli as they were being presented. A full stimulus list with the attention questions, can be downloaded from https://osf.io/f8grv/.

### Procedure and trial timing

The experiment was presented using E-prime 2.0 as illustrated in Figure 3. The task consisted of 270 trials divided into 9 blocks. In between each block, we offered the participants a break. Participants completed a practice block first to familiarise themselves with the paradigm.

**Figure 3.**
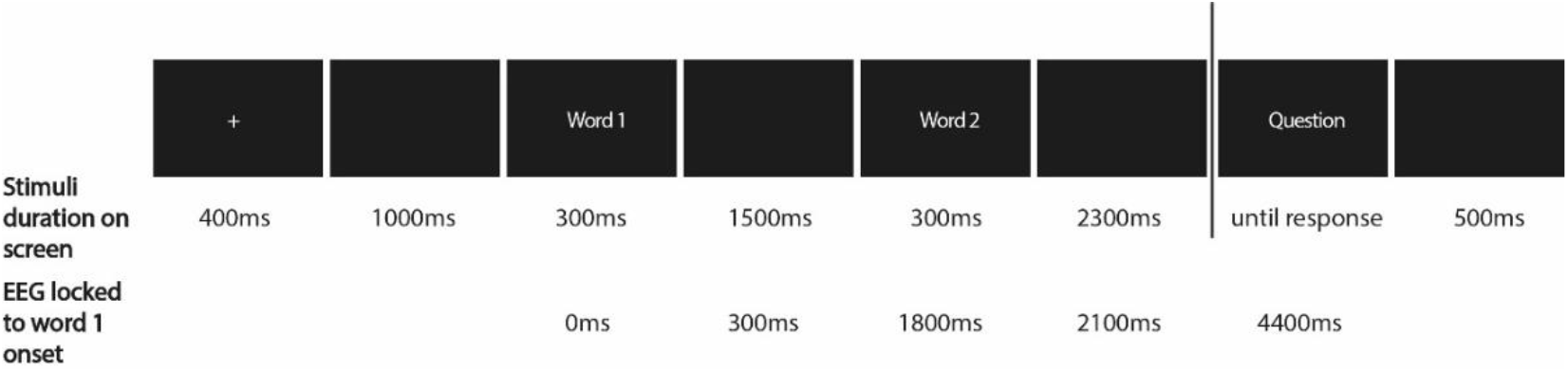
Trial presentation of the two-word phrase paradigm. The questions appeared in 22% of the trials.

### EEG recording

*cEEGrids* (Bleichner & Debener, 2017) were placed on skin around the ear including the mastoid bone, using double-sided adhesive. cEEGrids were thus located partly over the inferior temporal cortex. The use of a small amount of electrolyte enables low impedance electrode-skin contact. The right mastoid (R5) served as the ground while the left mastoid (L6) served as the reference during the online recordings (see Figure 1 for layout).

EEG for the *full-cap* group was recorded using Waveguard caps containing 64 cap-mounted Ag/AgCI electrodes (10-20 layout, including left and right mastoids), with online reference to the CPz channel. Horizontal eye movements were measured by two electrodes placed on the outer left and right canthi. Vertical eye movements were recorded by two electrodes placed above and below the right eye.

Both the 64-channel *full-cap* and *cEEGrids* signal was amplified with the ANTneuro EEGosports amplifier system and recorded using EEGo software (Advanced Neuro Technology). The signal was obtained using a 30Hz low-pass filter, a 0.05Hz (64-channel full-cap) / 0.3Hz (cEEGrid) high-pass filter and a 500Hz sampling rate. Impedances were kept below 20 kΩ.

### EEG analysis

The EEG pre-processing was performed using EEGLAB 14.1.2b (Delorme & Makeig, 2004) and Fieldtrip toolbox 2018-07-16 (Oostenveld et al., 2011). The data were epoched to the onset of the first word. The *cEEGrid* data were offline re-referenced to a linked mastoid where L6 and R6 served as the reference. The 64-channel *full-cap EEG* data were offline re-referenced to the average of all of the channels, where the mastoid and bipolar electrodes were excluded from the re-referencing. EEGLAB was used for manual inspection and rejection of trials with non-physiological artefacts. For the *full-cap EEG* data ocular artefacts were removed based on the scalp distribution and time-course, using an independent component analysis (ICA) extended algorithm in EEGLAB. For the *cEEGrid* data, ocular as well as heart-beat artefacts were removed through visual inspection of their ICA time course. There was no difference in the amount of ICA components rejected between the two groups (on average 2). Moreover, there was also no significant difference between the amount of trials rejected in the MCI (mean=28%, SD=16) and healthy elderly groups (mean=28%, SD =16).

The *cEEGrid* data were analysed in two ways. First, we performed analyses on the signal from the electrode L4. This electrode, which provided good signal quality across all participants and groups, was over the left temporal cortex which is a key region implicated in previous studies on language comprehension (Turken & Dronkers, 2011) and has been found to have reduced activation during word processing in MCI patients (Vandenbulcke et al., 2007).

In addition to the using the L4 electrode, we also used the data from all functioning electrodes in a participant by doing a principle component analysis (PCA) and conducting a time-frequency analysis on the first principle component. PCA is able to reduce the dimensions of the data through extracting orthogonal features (i.e components) of the data. The first component obtained in the PCA is the one that explains the most variance in the data.

Time-frequency representations (TFRs) of power were performed using Hanning tapers and the ‘mtmconvol’ method with a time window of 3 cycles per frequency of interest (DT = 3/f) for every trial, including 2Hz to 30Hz in steps of 1Hz. Changes in oscillatory power locked to the onset of the word were calculated in regard to the change in power from baseline. The data were baseline corrected to -600 to -100 ms prior to presentation of the first word.

To ensure that the observed oscillatory changes were not just the spectral representation of the ERPs, the ERP components were subtracted from the TFR (Mazaheri & Picton, 2005). The subtraction was achieved by first generating the time frequency decomposition of the ERP data for each condition and participant separately. Next, the time frequency power spectra (of the ERP) were subtracted from the time frequency power spectra of the EEG signal for each condition. The subsequent power changes in the time-frequency domain were used to generate time frequency power spectra differences between experimental conditions.

### Statistical analysis

The statistical differences of the experimental condition differences in the power changes in the time-frequency domain were assessed by using a two-tailed non-parametric cluster-based permutation test (using the FieldTrip toolbox) (Maris & Oostenveld, 2007).

For the 64-channel *full-cap EEG* data, the power of the frequencies of interest (theta: 4-7Hz, alpha: 8-14Hz, low-beta: 15-20Hz, high-beta: 20-25Hz) in each channel and time point from the onset of the first word to 1.4 seconds after the onset of the second word was subjected to a dependent samples t-test when comparing conditions. Likewise for the between group analysis, the power difference between conditions for each group was subjected to a independent sample t-test. Next, neighbouring electrodes (minimum of 2) and adjacent time points were clustered together if their t-value exceeded the threshold of P < 0.05 (two-tailed). The statistical significance of each cluster was assessed through randomly shuffling condition labels 1000 times and looking at the distribution of the t-values of the random clusters. Here a Monte Carlo derived P-value can be obtained through calculating the number of times the t-statistics in the shuffled distribution was higher than the original t-statistic derived by contrasting conditions. The cluster-level statistics were calculated by taking the sum of the t-values within every cluster, with the test statistic being the maximum of the cluster-level summed t-values (i.e ‘maxsum’ option in Fieldtrip).

We amended our approach to suit the analysis of the *cEEGrid* data. Given that only 1 channel of the cEEGrid data was analysed, we clustered the data across time-frequency tiles rather than clustering across channels, if their t-value exceeded the threshold of P < 0.05 (two-tailed). This reduction in dimensions allowed us to assess the differences in time-frequency representations between the conditions, without relying on pre-defined frequency bands of interest (similar to Segaert et al, 2018). Power was subjected to a dependent samples t-test comparing conditions from the onset of the first word to the onset of the second word (1.8 seconds later), and from the onset of the second word until 1.4 seconds after the onset of the this word. Likewise for the between group analysis the power difference between conditions for each group was subjected to a independent sample-test. Finally, in-line with the full-cap data EEG data, the cluster-level statistics were calculated by taking the sum of the t-values within every cluster, with the test statistic being the maximum of the cluster-level summed t-values (i.e ‘maxsum’ option in Fieldtrip).

### Assessing the diagnostic accuracy EEG signatures

We used the receiver operative characteristic (ROC) curve (Zou, O’Malley, & Mauri, 2007) to assess the sensitivity and specificity of single-word retrieval and binding signatures, detected using the cEEGRIDS, in distinguishing MCI patients from the healthy elderly. The ROC curve was estimated by varying the threshold of alpha/beta power in a certain range (defined by the difference between conditions *across participants*) and then calculating the sensitivity (true positive rate) and specificity (true negative rate). The ROC curve is a plot of sensitivity on the y axis against (1-specificity) on the x axis for varying values of the single-word retrieval and binding signatures. Sensitivity refers to the true positive rate (i.e. ability to correctly identify an MCI patient) and specificity refers to true negative rate (i.e. ability to correctly identify a healthy control). The Area Under the ROC curve (AUC) is able to provide an overall summary of diagnostic accuracy of detecting MCI patients. An AUC of 0.5 corresponds to a random chance of the signatures in classifying MCI patients from the healthy elderly, while a 1.0 represents perfect accuracy. We conducted our ROC analysis in SPSS 27.01.01 with the distribution assumption being nonparametric.

## Results

### Aberrant oscillatory power modulations observed during single-word retrieval in older adults with MCI compared to healthy controls

First, we qualitatively look at the cEEGrid results for each individual condition (for word 1) in Figure 4, to determine if the signal is comparable to what one would expect to see for full-cap data in response to words. In our cEEGrid electrode of interest, L4, for both real words and letter strings, the onset of the word induced an increase in theta (4-7Hz) power, followed by a suppression of alpha (8-14Hz) power, prior to an alpha power rebound. The pattern was more clearly visible for the healthy controls (top row of Figure 4) than the MCI patients (bottom row of Figure 4). We will return to the group comparisons later. First we focus on the pattern we see across conditions and groups, which indeed is consistent with previous studies investigating word processing using full-cap EEG, with a theta power increase visible most clearly around 0.2 sec post word onset (e.g. Bastiaansen et al., 2008; Bastiaansen et al., 2005) and a later alpha power suppression at around 0.4 sec post word onset (Davidson & Indefrey, 2007). The theta increase has been related to the processing of word forms (Bastiaansen et al., 2008; Bastiaansen et al., 2005) while the later alpha suppression at posterior sites has been associated with further post-perceptual processing of sensory information (Pfurtscheller, 2001) and allocation of resources according to processing demands (Van Diepen, Foxe, & Mazaheri, 2019). The similar patterns of oscillatory power modulations for word processing observed using the cEEGrids and the full-cap EEG suggest that they are picking up comparable signals.

**Figure 4.**
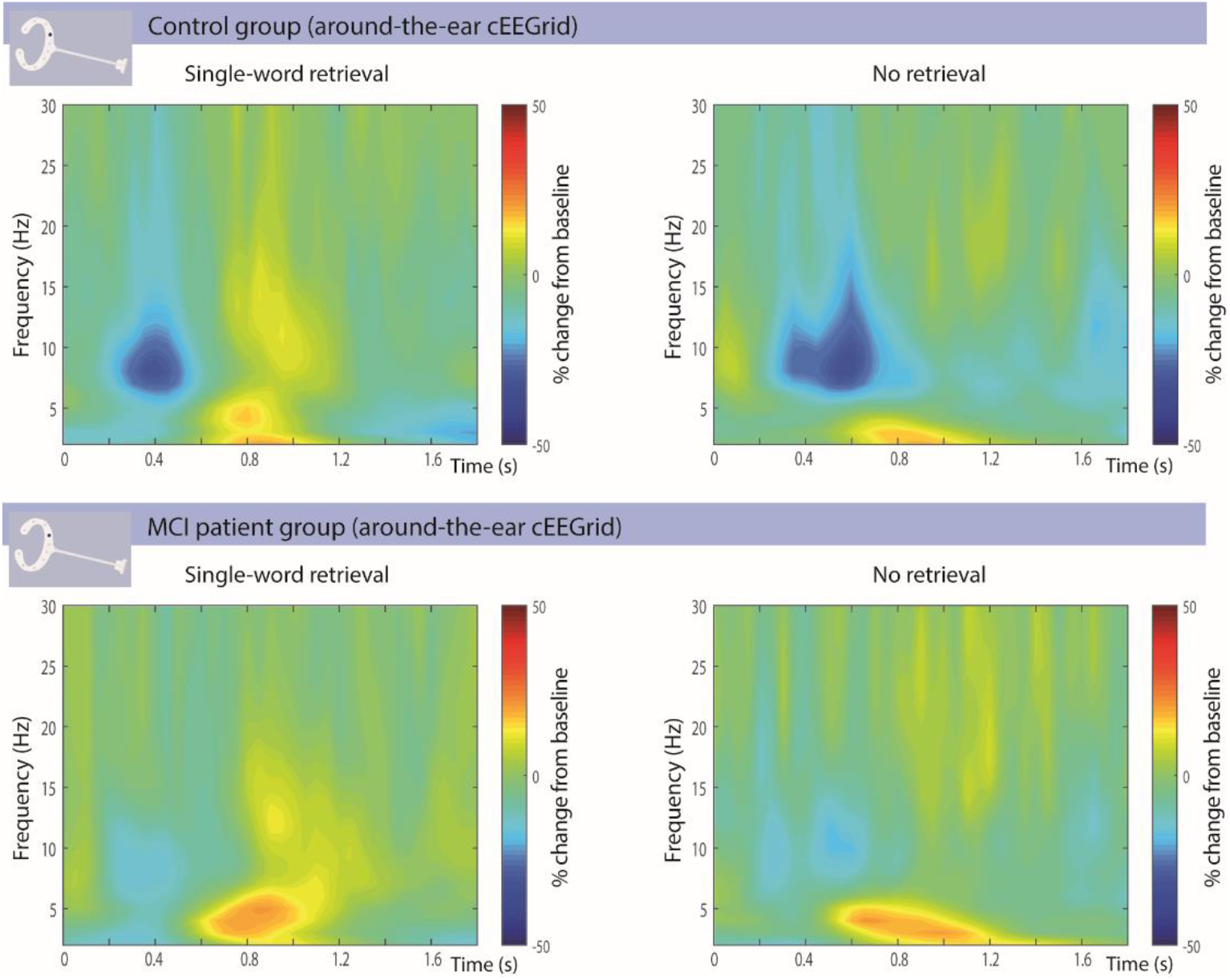
Time-frequency representations (TFRs) of power locked to the onset of the 1^st^ word for single-word retrieval of lexico-semantic properties (e.g. swift) and no retrieval (e.g. swrfeq). The TFRs are expressed as a percentage change from baseline (prior to onset of 1^st^ word) for cEEGrid electrode L4 (i.e. left-ear, coloured position in the depicted cEEGrid). Oscillatory changes induced by word-onset are illustrated for the healthy older adult controls (top row) and the MCI patients (bottom row). The word onset generated an increase in theta (4-7Hz) power, followed by a suppression of alpha (8-14Hz) power, and in turn followed by an alpha power rebound.

Next we turn to the condition differences. The differences in oscillatory EEG power between the single-word retrieval and no retrieval conditions serve as a signature for the lexico-semantic single-word retrieval effect. This condition difference is illustrated for the full-cap EEG data (top row) alongside the around-the-ear cEEGrid data (middle and bottom row) in Figure 5.

**Figure 5.**
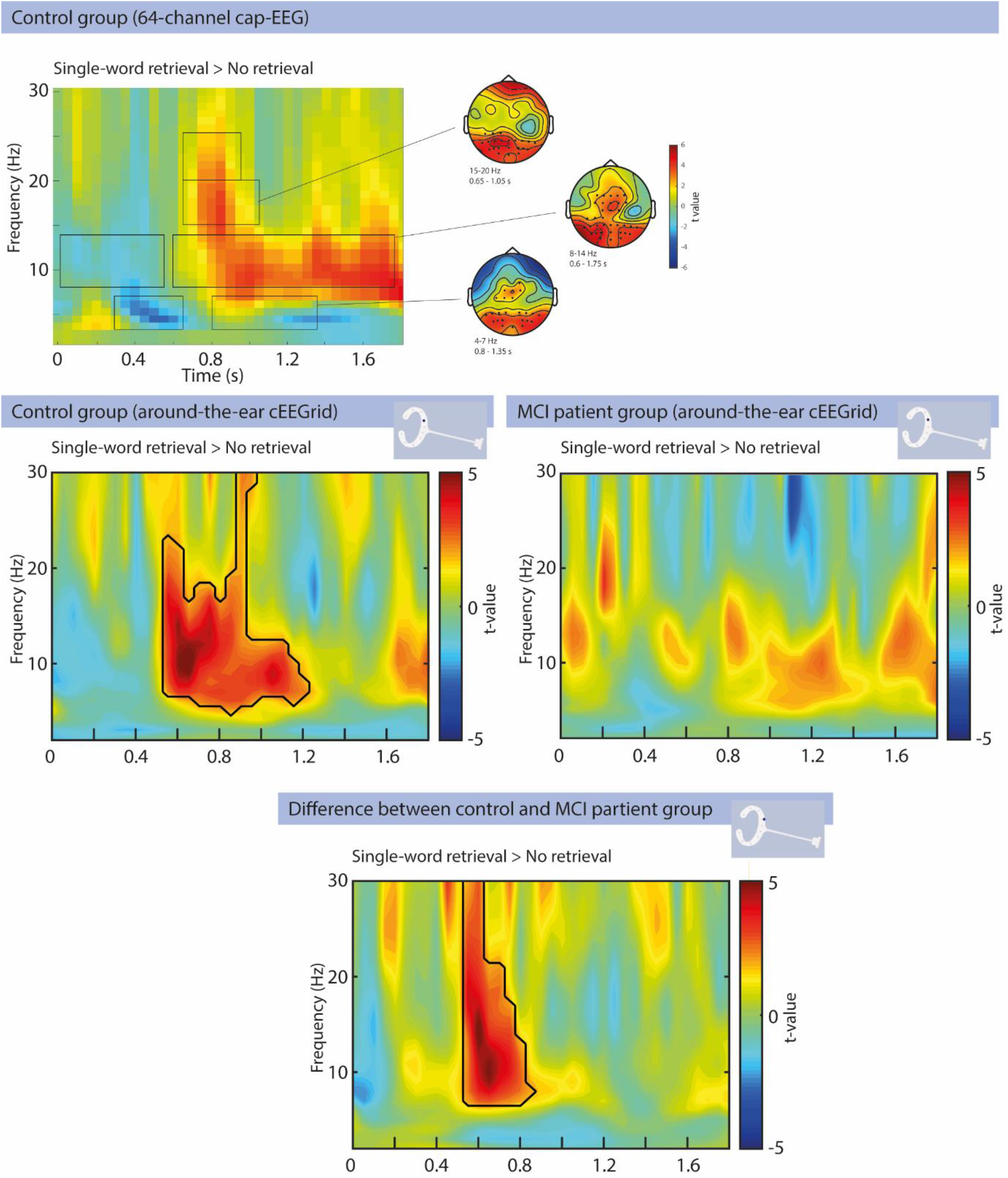
Oscillatory changes related to single-word retrieval of lexico-semantic properties in the 64 channel full-cap as well as the cEEGrid setup. The black lines contours inside the TFRs highlight the significant differences between single-word retrieval versus no retrieval conditions (swift vs. swrfeq) (p<0.05). The top row depicts the single-word retrieval condition effects in the full-cap EEG TFRs (averaged across all electrodes) for a group of healthy older adults, alongside (for selected effects) headplots with dots denoting clusters representing significant differences (based on data from Markiewicz, Segaert, Mazaheri, submitted). The middle row illustrates the cEEGrid (channel ‘L4’) single-word retrieval TFRs for the healthy older adult controls and MCI patients. The healthy controls had significant increase in the theta/alpha/beta power around 0.5-1.2 s after word-onset during single word lexical-retrieval (p<0.002). This effect did not reach significance in the MCI patients. The bottom row depicts the group comparison between healthy older adult controls and MCI patients for the cEEGrid data. The healthy controls had significantly greater alpha/beta activity around 0.5 -0.8 s after word-onset than the MCI patients during word-retrieval (p<.004).

We first describe the condition effects in a full-cap control group dataset, in order to later evaluate whether the cEEGrid condition effects in the control group are comparable. We found significant effects in pre-defined frequency bands, in theta (4-7Hz), alpha (8-14Hz), low-beta (15-20Hz) and high-beta (20-25Hz). In healthy older adults using the 64 channel EEG setup, single-word retrieval compared to the no retrieval condition showed a smaller theta increase (p = .002), corresponding to a cluster from 0.3 to 0.65 sec post word one onset maximal over right occipital and left central electrodes. There was also a greater theta increase (p = .004), corresponding to a cluster from 0.8 to 1.35sec and maximal over bilateral occipital and central channels. Furthermore, the retrieval condition elicited greater alpha suppression (p = .012), corresponding to a cluster from 0 to 0.55sec maximal over the bilateral parietal-central electrodes. This was followed by a greater alpha rebound (p<.002), corresponding to a cluser from 0.6 to 1.75sec maximal over the occipital and central electrodes. Similarly, there was greater low-beta power in the retrieval compared to the no retrieval condition (p < .002), corresponding to a cluster from 0.65 to 1.05sec over the occipital electrodes. Finally, we also observed greater high-beta power in the retrieval (compared to no retrieval) condition (p = .006), corresponding to a cluster from 0.65 to 0.95sec and maximal over the occipital channels.

In cEEGrid data we did not pre-define the frequency bands. In the healthy control group using the cEEGrids, the single-word retrieval effect as revealed in the L4 electrode was highly similar to the full-cap EEG data. For the cEEGrid data, we observed a significant condition difference (p<0.002) showing great power in the retrieval (compared to no retrieval) condition, in a cluster corresponding to ∼0.5 sec to ∼1.2 sec post-word onset and spanning across 5-30Hz range (corresponding to the theta, alpha and beta range). xInterestingly, within this time window we observed single-word retrieval condition differences in the full-cap EEG setup as well, which corresponded to clusters which were spatially maximal over left occipito-temporal electrodes (see illustrated topographies in top right of Figure 5). However we also note that there are effects observed in the full-cap data which were not observed in the c-EEGrid data, which could be due to the cEEGrids being blind to the sources producing these signals or due to the signal to noise not being at a level needed to detect them.

As can be seen in Figure 5, the single-word retrieval effect for the MCI patients was much less pronounced than for the healthy controls. For the MCI patients, the cEEGrid data do not reveal a significant single-word retrieval condition difference. Indeed, the comparison between the cEEGrid healthy control and the cEEGgrid MCI group reveals a statistical difference between the groups indicating the single-word retrieval condition effect is larger for the healthy older adult controls than the MCI patients (p<.004), in cluster corresponding to 0.5 sec to ∼0.8 sec and extending across 8-25Hz (corresponding to the alpha, low-beta and high-beta range).

In sum, following the typical post-word theta increase and alpha suppression effect, in the healthy control group we observed a clear rebound in the alpha and beta range when successful single-word retrieval of lexico-semantic properties is completed (compared to no successful retrieval of lexico-semantic properties). This rebound signature of successful retrieval completion was absent in the MCI group. These findings furthermore demonstrate that cEEGrids can reveal the expected oscillatory power modulations for single-word retrieval in healthy older adult controls, alongside aberrant oscillatory power modulations for single-word retrieval in older adults with MCI.

### Aberrant oscillatory power modulations supporting binding in older adults with MCI compared to healthy controls

The time-frequency representations of the cEEGrid data (‘L4’) locked to the onset of the 2^nd^ word in the binding and no binding context (i.e. individual conditions) can be seen in Figure 6. Similar to the onset of the 1^st^ word, the 2^nd^ word induced a transient increase in theta activity as well as an alpha suppression, followed by a rebound. This again suggests that the cEEGrid data pick up activity comparable to full-cap EEG.

**Figure 6.**
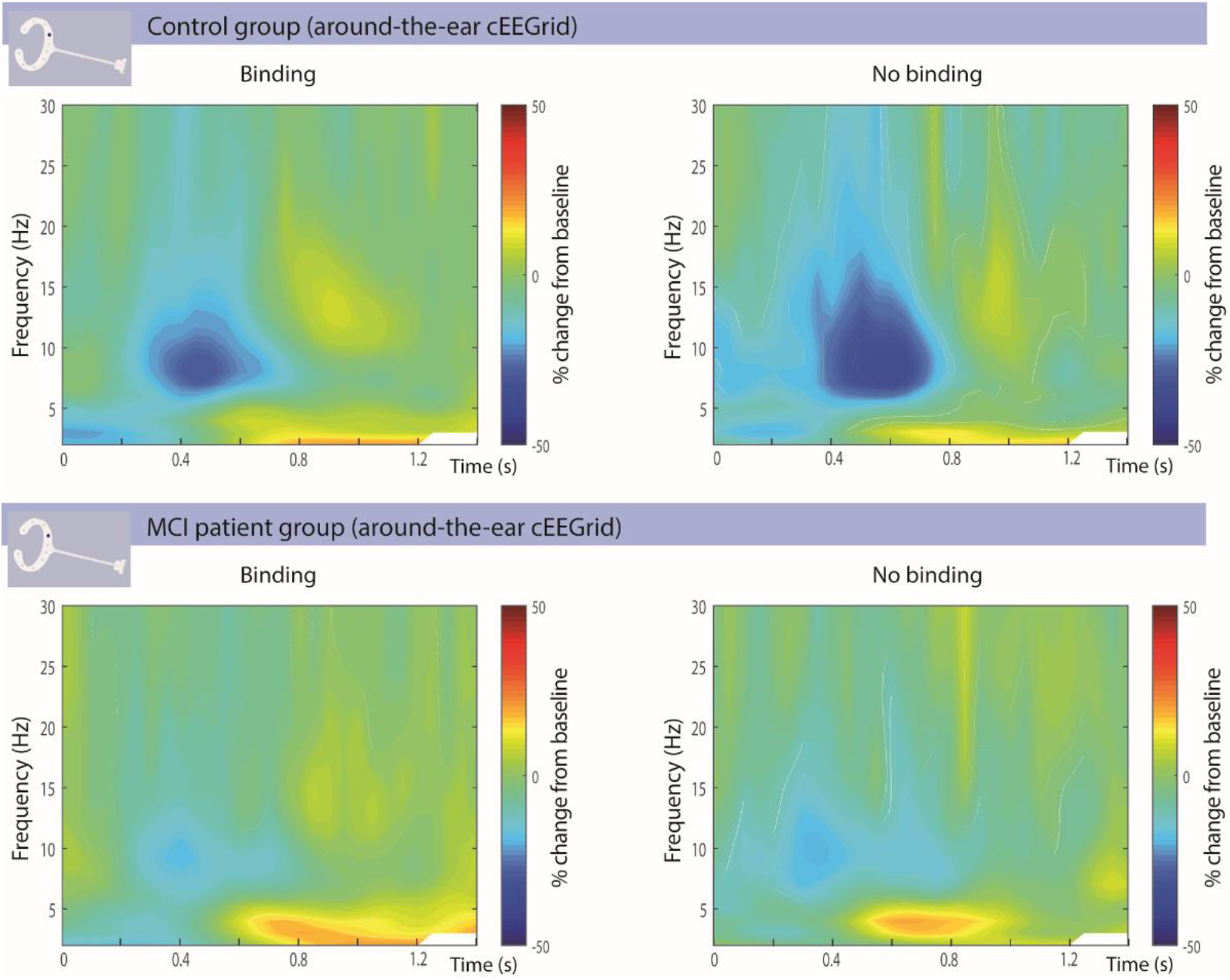
TFRs of power locked to the onset of the 2^nd^ word in a word pair, in a binding context (e.g. swift *horse*) and no binding context (e.g. swrfeq *horse*). The TFRs are expressed as a percentage change from baseline (prior to onset of 1^st^ word) for cEEGrid electrode L4 (i.e. left-ear, coloured position in the depicted cEEGrid). Similar to the 1^st^ word, the onset of the 2^nd^ word, irrespective of binding context, generated an increase in theta (4-7Hz) power, followed by a suppression of alpha (8-14Hz) power, and in turn followed by an alpha power rebound.

Figure 7 depicts the statistical comparison for the binding vs. no binding conditions (i.e. condition difference, or, the binding effect) for the full-cap EEG data alongside the cEEGrid data, for MCI patients and healthy controls.

**Figure 7.**
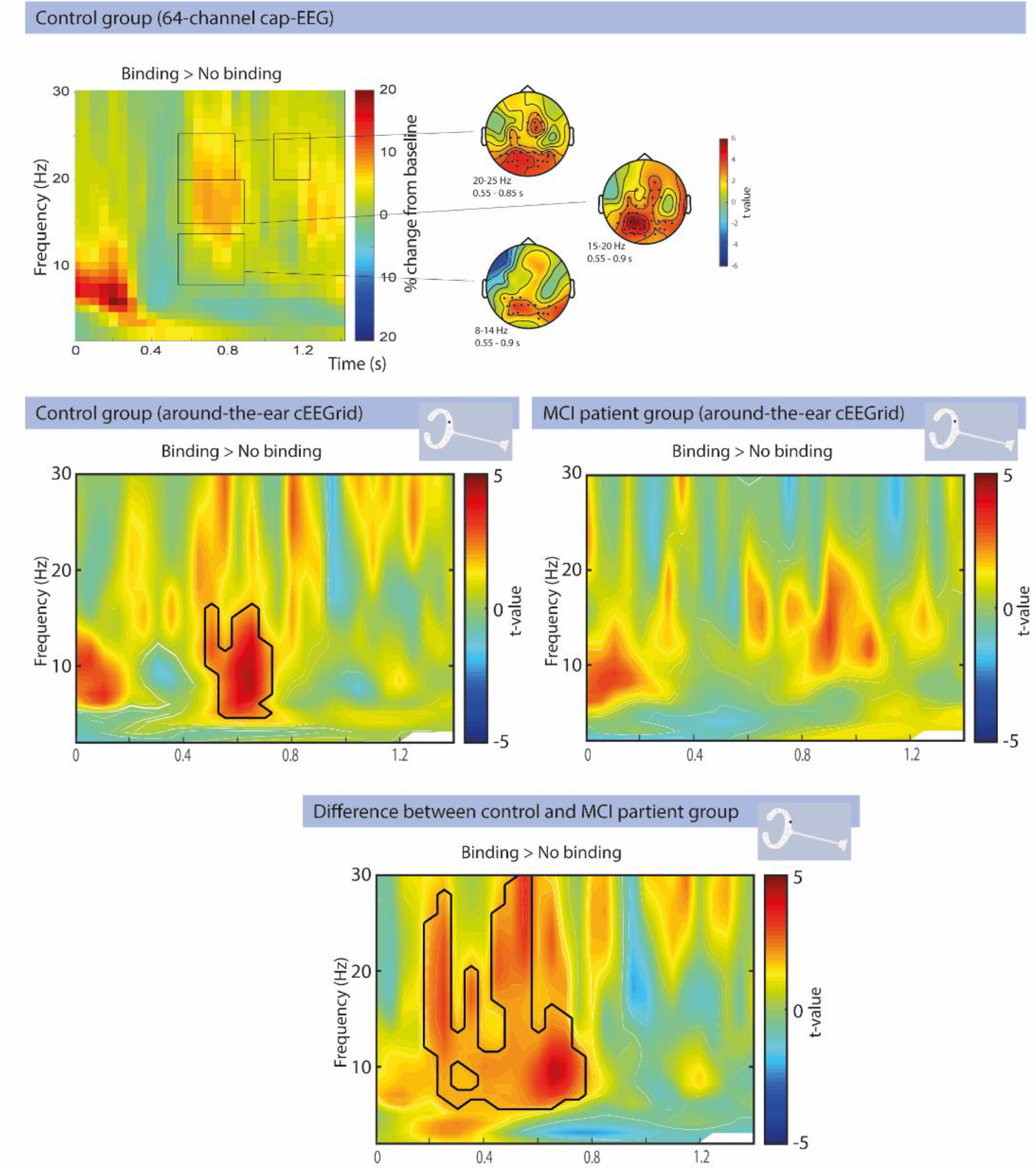
Oscillatory changes related to the binding condition difference, in the 64 channel full-cap as well as the cEEGrid setup. The black lines inside the TFRs highlight the significant (p<0.05) condition differences (i.e. binding compared to no binding). The top row depicts the binding effects in the full-cap EEG TFRs (averaged across all electrodes) for a group of healthy older adults, alongside (for selected condition effects) headplots with dots denoting clusters representing significant differences (based on data from Markiewicz, Segaert, Mazaheri, submitted). The middle row illustrates the cEEGrid (channel ‘L4’) binding effect TFRs for the healthy older adult controls and MCI patients. The healthy controls had a significant increase in alpha/beta power around 0.5 to 0.7 s during binding (p<0.03). This effect was not observed in the MCI patients. The bottom row depicts the comparison of oscillatory activity for the binding effect between healthy older adult controls and MCI patients in L4. The healthy older adults had significantly greater alpha/beta activity than the MCI patients (p<0.002) around 0.2-0.7 s after the onset of the word in the binding condition.

We first describe the condition effects in a full-cap control group dataset, in order to later on evaluate whether the cEEGrid condition effects in the control group are similar. In the full-cap data, for the binding compared to the no binding condition, there was reduced alpha suppression (p = .038), corresponding to a cluster from 0.55 to 0.9 sec and maximal over occipital and parietal electrodes. This condition difference extended into the low-beta (p = .002) and high-beta range (p = .002), corresponding to clusters from approximatly 0.55 to 0.9 sec and maximal over occipital and parietal electrodes. There was also a later condition effect in the high-beta range for the binding compared to the no binding condition (p = .044), corresponding to a cluster from 1.05 to 1.25 sec over occipito-central channels.

The cEEGrid data (electrode L4) for the healthy older adult controls revealed a highly similar binding effect to that observed in the full-cap EEG data. For the binding compared to the no binding condition in the control group, there was a reduced suppression in the alpha and low-beta range (p<.03), correspondings to a cluster from ∼0.5 sec to ∼0.7 sec.

As can be seen in Figure 6, the binding and no binding condition look highly similar to eachother within the MCI group. Indeed, we did not observe any significant condition differences (binding vs. no binding) in post-word oscillatory power in the MCI patients (Figure 7). A comparison between the cEEGrid healthy control group and the cEEGgrid MCI group (bottom row of Figure 7) confirms there is a statistical difference between the groups for the binding condition effect (p<.002), corresponding to a cluster from ∼0.2 sec to ∼0.7 sec extending over the alpha, low-beta and high-beta range.

In summary, in the healthy older adult control group there is a clear binding signature (i.e. less alpha and beta suppression for words in a binding compared to no binding context). This signature was absent in the MCI group. Together these findings show that cEEGrids can reveal the expected oscillatory power modulations for binding in healthy older adult controls, and that these signatures are significantly attenuated in older adults with MCI.

### Automated approach using the first principle component of the all cEEGrid channels revealed similar findings to the pre-defined region of interest approach (i.e. L4)

Similar to the results in the previous section, we found that the healthy older adult controls had a single-word retrieval effect (p<0.001, corresponding to a cluster from ∼.5 sec to ∼1.1 sec) extending over the alpha and low-beta range (see Figure 8), whereas there was no effect for the MCI group. As such there was a statistical difference between the groups (p<.01, corresponding to a cluster from ∼.5 sec to ∼8 sec). These results are highly similar to the results obtained when analysing electrode position L4.

**Figure 8.**
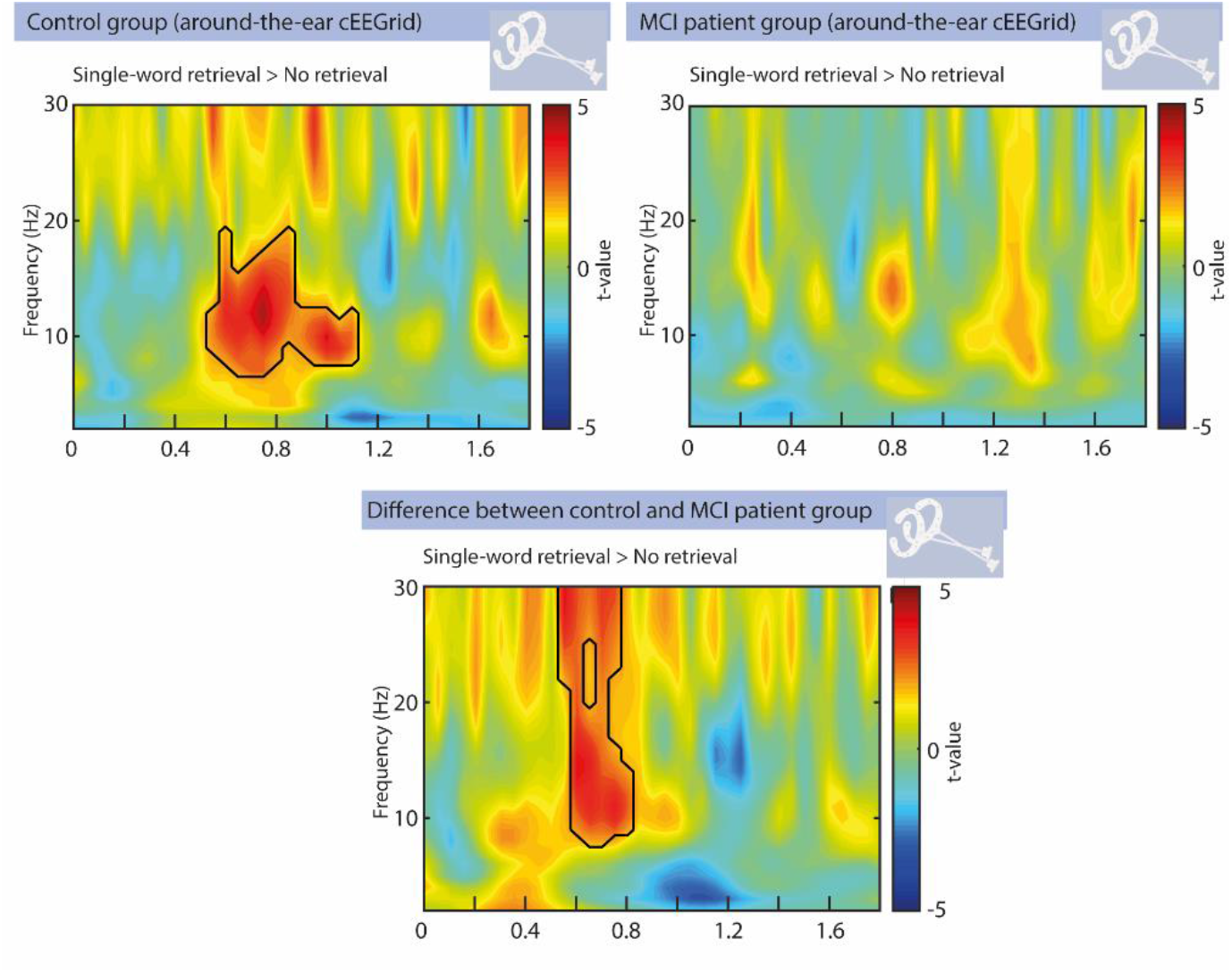
The single-word retrieval cEEGrid effects (i.e. single-word retrieval > no retrieval) for controls, the MCI patients and the group comparison, using an alternative way of analysing the cEEGrid data. Rather than using electrode position L4 for all participants (as was done for the results depicted in Figure 5), here we analysed the first principle component of all the cEEGrid electrodes in the participants. Top-left: healthy older adult controls had a significant increase in alpha/beta activity from 0.5-1.1 sec after the onset of the first word during single-word retrieval (p<0.001). Top-right: There were no significant time-frequency clusters during single-word retrieval found in the MCI group. Bottom: the healthy controls had significantly greater alpha/beta activity ∼0.5-0.8 s than MCI patients (p<0.01) after word onset, during single word retrieval.

For binding (see Figure 9), while qualitatively similar to the results obtained using the L4 location, the condition effect did not reach significance in the control group. The healthy controls nevertheless had a larger alpha/beta activity 0.4-0.8 s effect after the onset of the word than the MCI patients (p<0.005).

**Figure 9.**
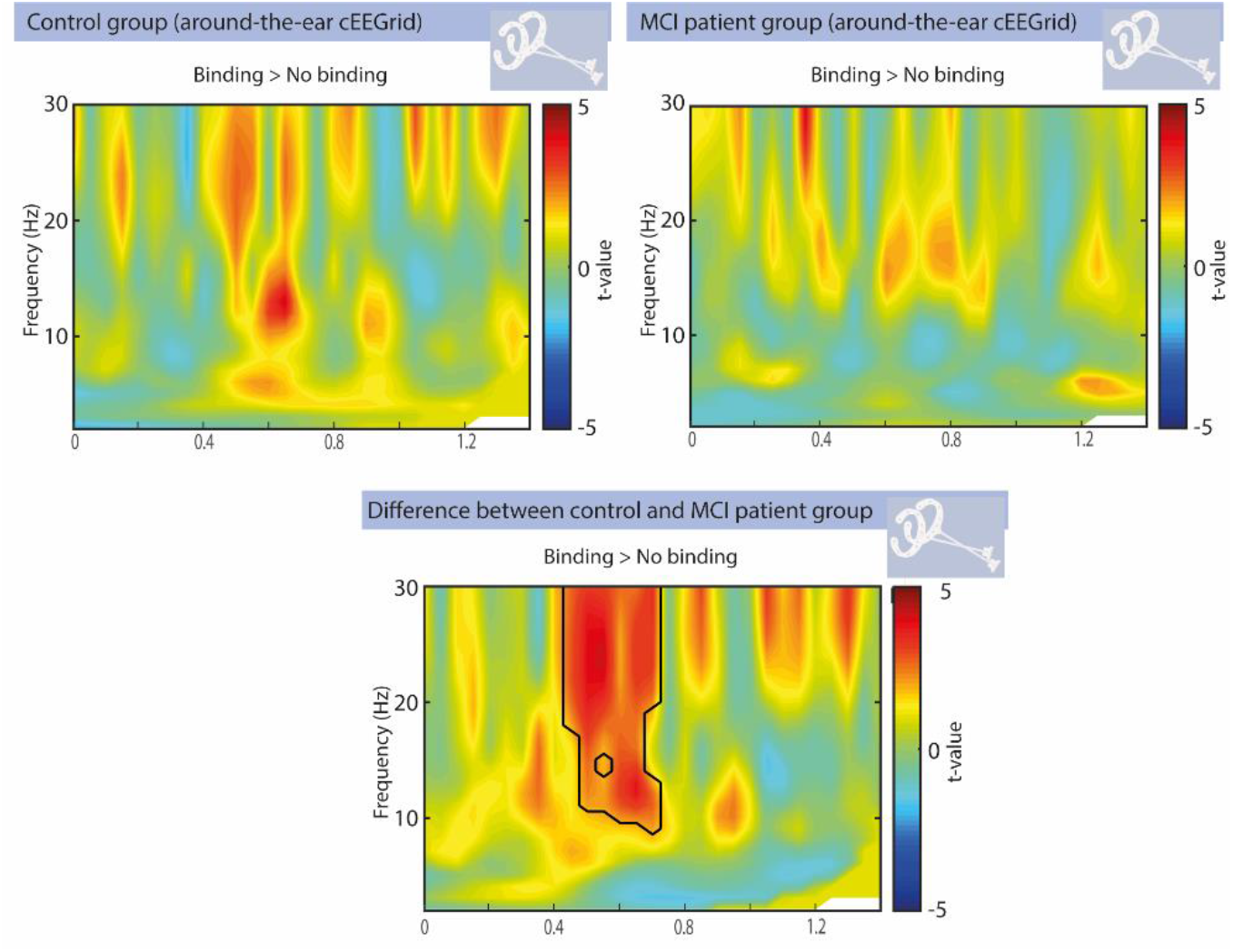
TFRs of power in the binding versus no binding condition, for the first principle component of all cEEGrid electrodes. While qualitatively similar to the results obtained using the L4 location, the condition effect did not reach significance in the control group. However, the controls still exhibited significantly greater alpha/beta activity ∼0.4-0.8 s after the onset of the word (p<0.005) for binding compared to the MCI patients.

### Sensitivity and specificity of single-word retrieval and multi-word binding EEG markers in differentiating MCI patients from healthy controls

Finally we set out to investigate the diagnostic potential of the single-word retrieval and multi-word binding signatures in detecting MCI patients from controls. The receiver operating characteristic curve (ROC) with the area under the curve (AUC) of retrieval and binding signatures can be seen in Figure 10. Here the sensitivity refers to the true positive rate (i.e. ability to correctly identify an MCI patient) and specificity refers to true negative rate (i.e. ability to correctly identify a healthy control). For the single-word retrieval signatures the AUC was 0.902 (SD=0.047, p<0.001, 95% CI: 0.81-0.993), while for the binding signatures the AUC was 0.866 (SD=.059, p<0.001, 95% CI: 0.75-0.98). While a larger sample size and cross-validation is needed to make any definitive claims about the diagnostic capabilities of these EEG markers, these preliminary AUROC results do suggest that these signals could potentially have excellent classification ability. Note that the classification ability here refers to the distinction between MCI patients and healthy controls, with the MCI cohort in this study being a heterogenuous sample.

**Figure 10.**
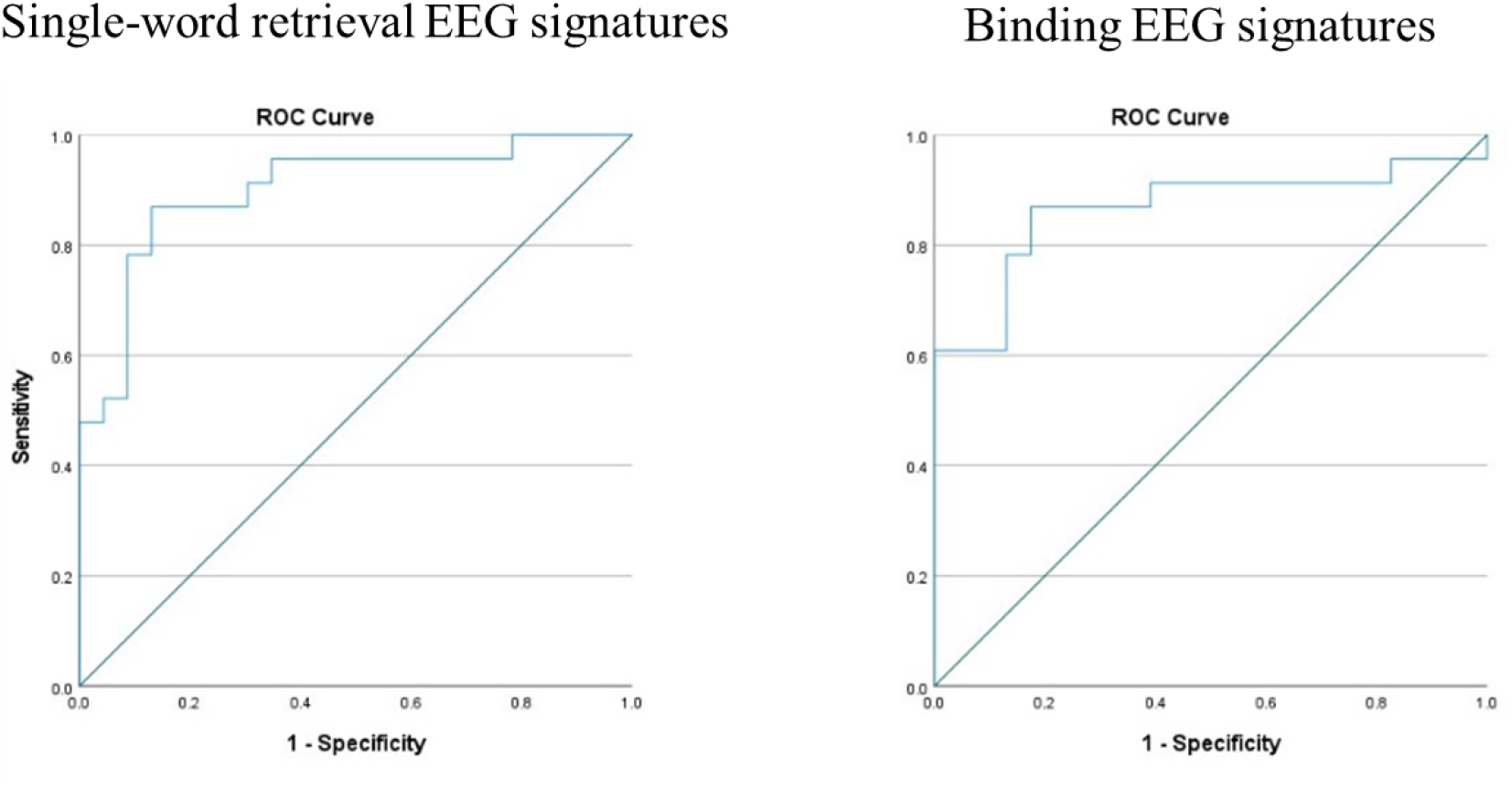
ROC curves for the single-word retrieval and multi-word binding signatures in distinguishing MCI patients from the healthy controls. For the retrieval signatures the AUC was 0.902 while for the binding signatures the AUC was 0.866 suggesting they have excellent to outstanding classification ability.

## Discussion

In the current study, we investigated if it was possible to detect subtle deficits during language comprehension in MCI patients using around-the-ear electrodes (cEEGrids). Language comprehension was tested in a two-word phrase paradigm which included a single-word retrieval manipulation (e.g. *swrfeq* versus *swift*) and a binding manipulation (e.g. *horse* preceded by *swift* versus preceded by *swrfeq*). Our conclusions are as follows. *First*, in the healthy control group, we observed, following a typical theta increase and alpha suppression effect, a clear rebound in the alpha/beta range when successful single-word retrieval was completed (compared to no single-word retrieval). This signature of successful single-word retrieval completion was absent in the MCI group. Our findings on word retrieval impairments in MCI are in line with previous findings showing deficits in this process (Mazaheri et al., 2018; Olichney et al., 2008; Taler & Jarema, 2006; Vandenbulcke et al., 2007). *Second*, MCI patients do not only have impairments for single-word processing, but also for multi-word binding combinatorics, i.e. building a meaning representation for multiple words. In the healthy control group there was a clear binding signature in the alpha and beta range (i.e. a reduced suppression effect in the alpha and beta range for the binding compared to the no binding condition) which was absent in the MCI group. This is a novel finding in the MCI literature, since previous studies have focused on single-word processing. *Third*, the sensitivity and specificity of the electrophysiological single-word retrieval signature as well as the binding signature for differentiating between MCI patients and controls was excellent. *Last*, we found that the cEEGrids can identify language comprehension signatures which are comparable to those observed with full-cap EEG set-ups, suggesting they are a sensitive tool to pick up subtle language impairments in clinical groups.

### Impairments in single-word retrieval for MCI patients

For the single-word retrieval effect, we observed significant differences in the alpha/beta rebound between the MCI patients and controls. In the healthy older adult control group, following the typical post-word theta increase and alpha suppression effect, we observed a clear rebound in the alpha and beta range when successful single-word retrieval was completed (compared to no single-word retrieval). This signature of successful single-word retrieval completion was absent in the MCI group. In line with previous work the alpha/beta rebound could be viewed to reflect suppression of further processing that could interfere with the encoded sensory information (Bonnefond & Jensen, 2012). This would in turn suggest that the MCI patients have an impairment in this gating mechanism.

A number of previous studies have observed an early word-induced (∼250-350 ms post-word) increase in theta activity, reflecting the processing of word forms (Bastiaansen et al., 2005; Bastiaansen et al., 2008; Hagoort et al., 2004). We previously found this theta increase to be significantly attenuated in MCI patients who would go on to develop Alzheimer’s disease within 3 years, relative to non-convertors and healthy elderly controls (Mazaheri et al., 2018). In our previous study, there was no difference in this early theta effect however between MCI non-convertors and controls. In the current study, we did not observe a significant word-induced theta difference between the MCI patients and controls, suggesting that we are likely looking at a mixed pool of MCI non-convertors and prodromal AD patients.

### Impairments in multi-word binding operations for MCI patients

Our study extends on previous knowledge of language comprehension deficits in MCI, and shows for the first time that MCI patients have impairments in binding, a key process for the comprehension of multi-word utterances. While the healthy older adult control group showed a clear binding signature in the alpha/beta range (in line with previous oscillatory signatures observed for binding (Meyer, 2018; Prystauka & Lewis, 2019; Weiss & Mueller, 2012)(Markiewicz et al., submitted; Poulisse et al., 2020), the MCI group did not show this binding signature. This is a novel finding since previous work has focused on single-word processing deficits. Identifying sensitive measures for different language comprehension impairments in MCI can contribute to the implementation of a more diverse battery of cognitive tests for MCI patients, which could include single-word as well as multi-word language comprehension deficits.

### The potential of the cEEGrids as a clinical tool

While a larger sample size and cross-validation is needed to make any definitive claims about the diagnostic capabilities of the single-word retrieval and multi-word binding EEG signatures, our preliminary AUROC results do suggest that these signals could potentially have excellent classification ability in distinguishing MCI from healthy controls. Moreover, a novel aspect of the present work is the use of cEEGrids (Bleichner & Debener, 2017). This novel, unobtrusive, fast-to-apply electrode array could be instrumental in mapping language comprehension dysfunction in MCI patients. The current study presents converging evidence on the applicability and use of around-the-ear cEEGrids for mapping cognitive signatures (Bleichner & Debener, 2017).

Based on our experience in the current study, we believe that cEEGrids are an exciting avenue to conveniently and quickly acquire clinical EEG data. However we have come across a few issues that should be considered by researchers interested in using cEEGrids. We initially manufactured an adapter in-house to link the cEEGrids to our amplifier. This adapter often became loose when participants moved their heads resulting in signal drop out. This issue was largely remedied when we acquired a newly designed adapter (https://www.easycap.de/). However, not even this adapter was completely resilient to gross neck movements of participants. We advise future studies to consider patient movement and comfort when interfacing the cEEGrids with their amplifiers.

One other issue we feel is worth discussing is the choice of electrodes. We prioritized rapid application of the ceegrids, then reducing the impedance of the electrodes. This invariably led to electrodes near the face often having less contact/ poorer signal than other cEEGrids channels, particularly when the participants had facial hair. We had a prior region of interest for our study based on previous work (i.e. L4, see above), and we suggest that future work should consider a priori what brain areas are most relevant to their design and research question. Alternatively, using a feature extraction method such as PCA, we were still able to obtain meaningfull results from the wearable electrodes, without the need for pre-defined locations.

We note futhermore that the present cEEGrid study focused on language comprehension, which generates posterior scalp activity. Likewise, most previous cEEGrid studies investigated auditory processing (Bleichner & Debener, 2017; Bleichner, Mirkovic, & Debener, 2016; Debener, Emkes, De Vos, & Bleichner, 2015). Taken together with findings demonstrating that cEEGrids are best suited for recording activity from posterior scalp sites (Pacharra, Debener, & Wascher, 2017), it is likely that cEEGrids are not equally suited for all cognitive tasks.

### Conclusions

Using novel around-the-ear electrodes (cEEGrids) to assess language comprehension, the single-word retrieval and multi-word binding oscillatory power modulations observed in control participants were absent in MCI patients. These findings indicate that EEG can be used to deliver a neurophysiological correlate of language comprehension impairments in MCI, and that these impairments are not limited to the comprehension of single-words but also affect binding operations, which is essential to the comprehension of multi-word utterances.

## Data Availability

Anonymized data are available on request pending approval from ethics body.

## Acknowledgements

The authors would like to thank all the participants who contributed to this research.

## References

Ahmed, S., Arnold, R., Thompson, S. A., Graham, K. S., & Hodges, J. R. (2008). Naming of objects, faces and buildings in mild cognitive impairment. Cortex, 44(6), 746–752.

Alexopoulos, P., Grimmer, T., Perneczky, R., Domes, G., & Kurz, A. (2006). Progression to dementia in clinical subtypes of mild cognitive impairment. Dementia and geriatric cognitive disorders, 22(1), 27–34.

Bastiaansen, M., Oostenveld, R., Jensen, O., & Hagoort, P. (2008). I see what you mean: theta power increases are involved in the retrieval of lexical semantic information. Brain and Language, 106(1), 15–28.

Bastiaansen, M., Van Der Linden, M., Ter Keurs, M., Dijkstra, T., & Hagoort, P. (2005). Theta responses are involved in lexical—Semantic retrieval during language processing. Journal of Cognitive Neuroscience, 17(3), 530–541.

Bleichner, M. G., & Debener, S. (2017). Concealed, unobtrusive ear-centered EEG acquisition: cEEGrids for transparent EEG. Frontiers in Human Neuroscience, 11, 163.

Bleichner, M. G., Mirkovic, B., & Debener, S. (2016). Identifying auditory attention with ear-EEG: cEEGrid versus high-density cap-EEG comparison. Journal of neural engineering, 13(6), 066004.

Bonnefond, M., & Jensen, O. (2012). Alpha oscillations serve to protect working memory maintenance against anticipated distracters. Current biology, 22(20), 1969–1974.

Bozoki, A., Giordani, B., Heidebrink, J. L., Berent, S., & Foster, N. L. (2001). Mild cognitive impairments predict dementia in nondemented elderly patients with memory loss. Archives of Neurology, 58(3), 411–416.

Caramelli, P., Mansur, L., & Nitrini, R. (1998). Language and Communication Disorders in Dementia of the Alzheimer Type.

Davidson, D. J., & Indefrey, P. (2007). An inverse relation between event-related and time–frequency violation responses in sentence processing. Brain Research, 1158, 81–92.

Debener, S., Emkes, R., De Vos, M., & Bleichner, M. (2015). Unobtrusive ambulatory EEG using a smartphone and flexible printed electrodes around the ear. Scientific reports, 5, 16743.

Duong, A., Whitehead, V., Hanratty, K., & Chertkow, H. (2006). The nature of lexico-semantic processing deficits in mild cognitive impairment. Neuropsychologia, 44(10), 1928–1935.

Eyigoz, E., Mathur, S., Santamaria, M., Cecchi, G., & Naylor, M. (2020). Linguistic markers predict onset of Alzheimer’s disease. EClinicalMedicine, 28, 100583.

Foxe, J. J., Simpson, G. V., & Ahlfors, S. P. (1998). Parieto-occipital∼ 1 0Hz activity reflects anticipatory state of visual attention mechanisms. Neuroreport, 9(17), 3929–3933.

Garrett, M., Debener, S., & Verhulst, S. (2019). Acquisition of subcortical auditory potentials with around-the-ear cEEGrid technology in normal and hearing impaired listeners. Frontiers in neuroscience, 13, 730.

Gauthier, S., Reisberg, B., Zaudig, M., Petersen, R. C., Ritchie, K., Broich, K., … Chertkow, H. (2006). Mild cognitive impairment. The lancet, 367(9518), 1262–1270.

Henry, J. D., Crawford, J. R., & Phillips, L. H. (2004). Verbal fluency performance in dementia of the Alzheimer’s type: a meta-analysis. Neuropsychologia, 42(9), 1212–1222.

Markiewicz, R., Segaert, K., & Mazaheri, A. (submitted). How the healthy ageing brain supports semantic binding during language comprehension. Preprint available on bioRxiv. doi:https://doi.org/10.1101/2021.01.15.426707

Mazaheri, A., Segaert, K., Olichney, J., Yang, J.-C., Niu, Y.-Q., Shapiro, K., & Bowman, H. (2018). EEG oscillations during word processing predict MCI conversion to Alzheimer’s disease. NeuroImage: Clinical, 17, 188–197.

Mioshi, E., Dawson, K., Mitchell, J., Arnold, R., & Hodges, J. R. (2006). The Addenbrooke’s Cognitive Examination Revised (ACE-R): a brief cognitive test battery for dementia screening. Int J Geriatr Psychiatry, 21(11), 1078–1085. doi:10.1002/gps.1610

Mirkovic, B., Bleichner, M. G., De Vos, M., & Debener, S. (2016). Target speaker detection with concealed EEG around the ear. Frontiers in neuroscience, 10, 349.

Mitchell, A. J., & Shiri-Feshki, M. (2009). Rate of progression of mild cognitive impairment to dementia–meta-analysis of 41 robust inception cohort studies. Acta psychiatrica scandinavica, 119(4), 252–265.

O’Bryant, S. E., Humphreys, J. D., Smith, G. E., Ivnik, R. J., Graff-Radford, N. R., Petersen, R. C., & Lucas, J. A. (2008). Detecting Dementia With the Mini-Mental State Examination in Highly Educated Individuals. Archives of Neurology, 65(7), 963–967. doi:10.1001/archneur.65.7.963

Olichney, J. M., Taylor, J., Gatherwright, J., Salmon, D., Bressler, A., Kutas, M., & Iragui-Madoz, V. (2008). Patients with MCI and N400 or P600 abnormalities are at very high risk for conversion to dementia. Neurology, 70(19 Part 2), 1763–1770.

Pacharra, M., Debener, S., & Wascher, E. (2017). Concealed around-the-ear EEG captures cognitive processing in a visual simon task. Frontiers in Human Neuroscience, 11, 290.

Pfurtscheller, G. (2001). Functional brain imaging based on ERD/ERS. Vision research, 41(10-11), 1257–1260.

Portet, F., Ousset, P., Visser, P., Frisoni, G., Nobili, F., Scheltens, P., … Disease, M. W. G. o. t. E. C. o. A. s. (2006). Mild cognitive impairment (MCI) in medical practice: a critical review of the concept and new diagnostic procedure. Report of the MCI Working Group of the European Consortium on Alzheimer’s Disease. Journal of Neurology, Neurosurgery & Psychiatry, 77(6), 714–718.

Poulisse, C., Wheeldon, L., Limachya, R., Mazaheri, A., & Segaert, K. (2020). The oscillatory mechanisms associated with syntactic binding in healthy ageing. Neuropsychologia, 146, 107523.

Poulisse, C., Wheeldon, L., & Segaert, K. (2019). Evidence against preserved syntactic comprehension in healthy aging. Journal of Experimental Psychology: Learning, Memory and Cognition, 45(12), 2290–2308.

Roark, B., Mitchell, M., Hosom, J.-P., Hollingshead, K., & Kaye, J. (2011). Spoken language derived measures for detecting mild cognitive impairment. IEEE transactions on audio, speech, and language processing, 19(7), 2081–2090.

Sterr, A., Ebajemito, J. K., Mikkelsen, K. B., Bonmati-Carrion, M. A., Santhi, N., Della Monica, C., … Debener, S. (2018). Sleep EEG derived from behind-the-ear electrodes (cEEGrid) compared to standard polysomnography: A proof of concept study. Frontiers in Human Neuroscience, 12, 452.

Taler, V., & Jarema, G. (2006). On-line lexical processing in AD and MCI: An early measure of cognitive impairment? Journal of neurolinguistics, 19(1), 38–55.

Taler, V., & Phillips, N. A. (2008). Language performance in Alzheimer’s disease and mild cognitive impairment: a comparative review. Journal of clinical and experimental neuropsychology, 30(5), 501–556.

Turken, A., & Dronkers, N. (2011). The Neural Architecture of the Language Comprehension Network: Converging Evidence from Lesion and Connectivity Analyses. Frontiers in systems neuroscience, 5(1). doi:10.3389/fnsys.2011.00001

Van Diepen, R. M., Foxe, J. J., & Mazaheri, A. (2019). The functional role of alpha-band activity in attentional processing: the current zeitgeist and future outlook. Current opinion in psychology, 29, 229–238.

Vandenbulcke, M., Peeters, R., Dupont, P., Van Hecke, P., & Vandenberghe, R. (2007). Word reading and posterior temporal dysfunction in amnestic mild cognitive impairment. Cerebral Cortex, 17(3), 542–551.

Weiss, S., & Mueller, H. M. (2003). The contribution of EEG coherence to the investigation of language. Brain and language, 85(2), 325–343.

Zou, K. H., O’Malley, A. J., & Mauri, L. (2007). Receiver-operating characteristic analysis for evaluating diagnostic tests and predictive models. Circulation, 115(5), 654–657.

